# Utilization of a SARS-CoV-2 Variant Assay for the Rapid Differentiation of Omicron and Delta

**DOI:** 10.1101/2021.12.22.21268195

**Authors:** Nicholas Barasch, Jameel Iqbal, Marvin Coombs, Sofia Kazi, Jessica Wang-Rodriguez, Mark Holodniy, Michael Gelman

## Abstract

The emergence of the severe acute respiratory syndrome coronavirus 2 (SARS-CoV-2) Omicron variant (B.1.1.529), creates a diagnostic vacuum, since differentiation of Omicron from Delta relies on relatively slow next generation sequencing (NGS) technology delaying epidemiologic understanding and therapeutic intervention. The RUO SARS-CoV-2 Variant Set 1 Test (RSCov2V1) RT-PCR for detection of spike gene N501Y, E484K and del69-70 was designed to differentiate Alpha from Beta and Gamma variants. While Delta lacks these three variants, Omicron has the N501Y and del69-70 mutation. We submitted 88 samples for RSCov2V1 identifying 9 samples with the N501Y and del69-70 mutations while all other samples (79) were negative for all three variants. 9/9 samples with the del69-70 and N501Y were identified by NGS to be Omicron while 47/47 other samples assessed by NGS were confirmed to be Delta family variants. We demonstrate here that an immediately available RT-PCR assay for detection of spike gene N501Y and del69-70 can be utilized to rapidly differentiate Omicron from Delta variants in the proper epidemiologic context

## Introduction

The emergence of new variants of severe acute respiratory syndrome coronavirus 2 (SARS-CoV-2), such as the Omicron variant (B.1.1.529), creates a diagnostic vacuum, since differentiation of emerging variants from previously dominant variants relies on time-consuming, capacity-limited next generation sequencing (NGS). This delay sacrifices both real-time epidemiologic insight and the opportunity for early precision antibody therapy. *De novo* development of rapid real time (RT)-PCR assays to distinguish multiple variants also appears less desirable; since newer variants may have none of the hallmark mutations of older variants, such assays risk obsolescence. This was the fate of the commercial Roche RUO SARS-CoV-2 Variant Set 1 Test (RSCov2V1) for detection of spike gene N501Y, E484K and del 69-70 which was designed to differentiate Alpha, Beta, and Gamma variants from their ancestral lineage. From September to November 2021, over 6,000 SARS-CoV-2 specimens were sequenced by the Veterans Health Administration (VHA) SeqForce consortium, identifying only Delta and its sub-lineages, which lack these three mutations. As Delta dominated other variants, the RSCov2V1’s value was apparently lost. However, Omicron shares the N501Y and del69-70 mutations with Alpha (Figure),^1^ implying that assays such as RSCov2V1 can be repurposed to differentiate rapidly between Omicron and Delta. Here we demonstrate that the RSCov2V1 assay can rapidly differentiate Omicron from a background of Delta specimens.

**Figure.**
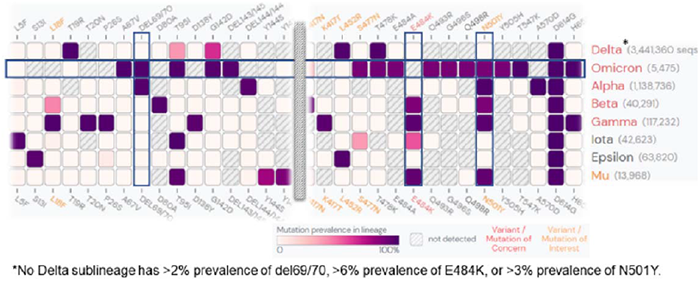
Target regions of RSCov2V1 as seen across multiple SARS-CoV-2 variants, including Omicron and Delta. Data source: GISAID. Visualization generated using outbreak.info.^1^

## Methods

Pharyngeal swabs in viral or universal transport media (VTM/UTM) collected at multiple VA hospitals were tested for SARS-CoV-2 using routine RT-PCR platforms at those sites. Positive samples with a cycle threshold (Ct) value of <32 were assayed using RSCov2V1. 9 specimens positive for any target, and a subset of 47 of 79 specimens negative for all targets (the limit of our sequencing capacity prior to this manuscript submission), were reflexed to SARS-CoV-2 NGS (see Supplementary Methods for details). Because these data were generated during operational validation of a laboratory-developed test for clinical use, this activity was determined by our IRB not to constitute research.

## Results

88 unique-patient samples, previously positive for SARS-CoV-2 by RT-PCR from multiple VA hospitals across New York, New Jersey, Pennsylvania, West Virginia, Washington DC, and Florida, were assayed by RSCov2V1. 9 samples were positive for N501Y and del69-70 and 9/9 were confirmed as the omicron variant by NGS. The remaining 79 samples were negative for all 3 targets by RSCov2V1; a subset of 47/79 was submitted for NGS, and 47/47 were found to be Delta-family lineages (Table).

**Table.**
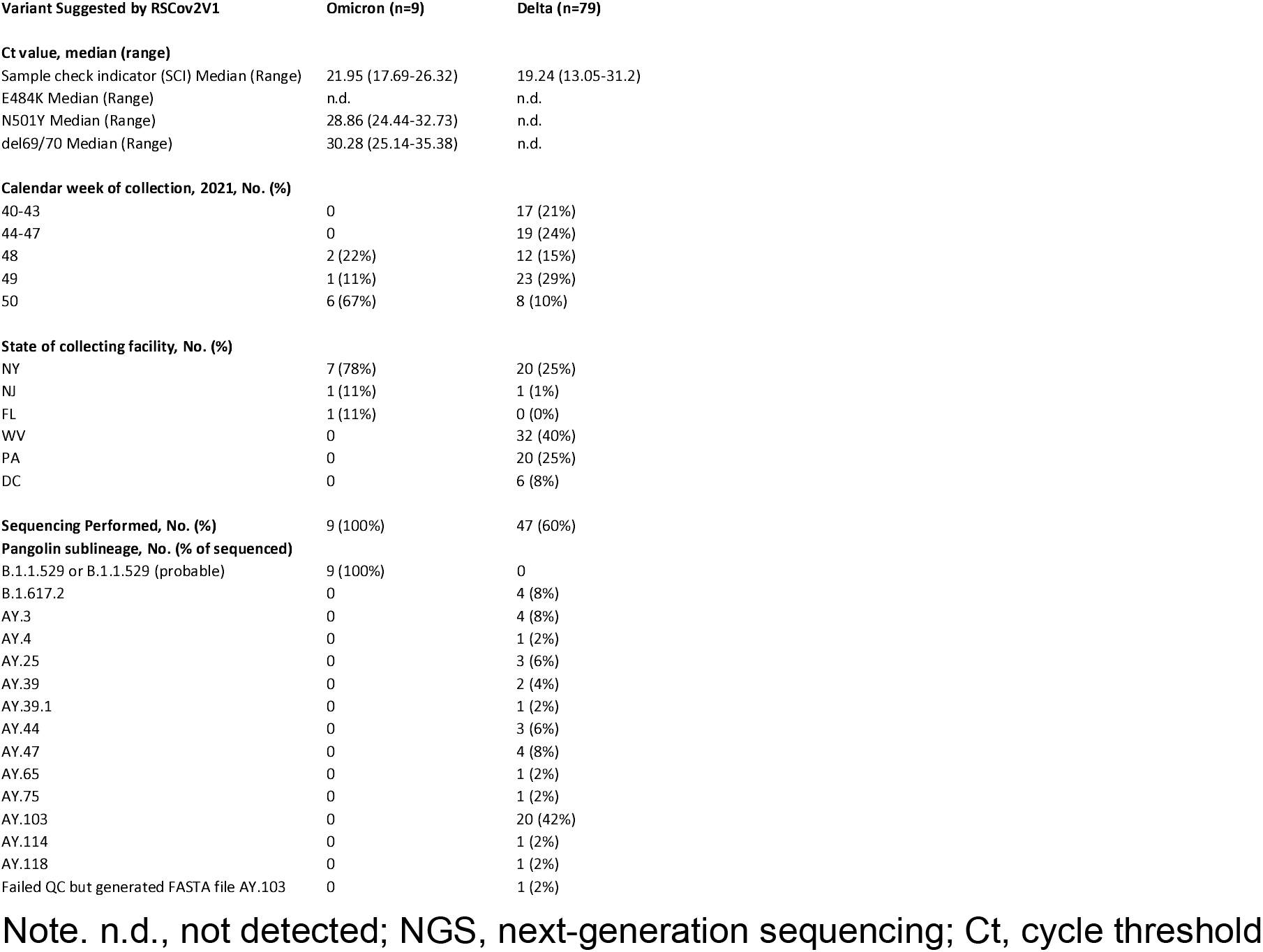
88 Samples Analyzed by RSCov2V1 to Differentiate Delta from Omicron.

## Discussion

As the proportion of Omicron increases in our region, we have been using RSCov2V1 as a screening tool to prioritize probable Omicron samples for subsequent NGS confirmation. Whereas NGS is relatively slow (taking up to 3 days), and requires highly trained personnel, by contrast, RSCov2V1 produces 93 results per 4-hour run with sample preparation. Current guidelines both recommend monoclonal antibody (mAb) therapy as early as possible after a positive viral test and predict that 2/3 FDA-authorized mAb therapies are unlikely to be active against Omicron.^2^ In situations where Delta and Omicron are both present in a population, variant RT-PCR allows for early precision mAb therapy, targeting limited supplies of sotrovimab to those with Omicron and extending the utility of other mAb therapies for remaining Delta. It also allows earlier and more complete capture of Omicron variant prevalence and more rapid generation of evidence about clinical features such as its disease severity. In addition, the insight into regional variant prevalence could enable timely allocation of different monoclonal therapies.

In summary, we demonstrate here that an immediately available RT-PCR assay can be utilized to rapidly differentiate Omicron from Delta variants. In future variant waves, similar strategies may be useful for detection of newer variants against a dominant background.

## Data Availability

All data produced in the present work are contained in the manuscript.

## Author Contributions

Nicholas Barasch MD Conception, Data Generation, Manuscript Generation, Review and Editing

Jameel Iqbal MD PhD Conception, Manuscript Generation, Review and Editing Marvin Coombs MPH MBA Data Generation, Manuscript review and editing Sofia Kazi MD Administrative support, Manuscript review and editing

Jessica Wang-Rodriguez MD Administrative support, Manuscript review and editing

Mark Holodniy MD Provided Samples, Manuscript Generation, Review and Editing

Michael Gelman MD PhD Conception, Manuscript Generation, Review and Editing

## The authors would like to acknowledge

Rozita Emami-Gorizi, Esthefany B Lizardo De Jimenez, Nissa Leal, Hame Balgobin and Carmen Salgado for rapid running and validation of the RSCov2V1, Idamary Perozo, Deaette Mclauren and Gerald Famby for development of our NGS platform and Carlos Campo for administrative support.

