## Supplemental Methods for "Utilization of a SARS-CoV-2 Variant Assay for the Rapid Differentiation of Omicron and Delta"

Supplementary Methods:

Funding/Project Development/Disclaimer: This work was conducted with Department of Veterans Affairs (VA) operational funding as part of standard assay validation for a laboratory-developed test for clinical use. All reagents were purchased at prices quoted to the VA prior to the emergence of Omicron. The assay manufacturer did not participate in the funding, design, conduct, or review of this work.

Specimens: Nasopharyngeal swab specimens were collected in viral/universal transport media (VTM/UTM) at VA medical centers in New York, New Jersey, Washington DC, Pennsylvania, West Virginia and Florida. SARS-CoV-2 detection was determined using various RT-PCR platforms at these sites. Positive SARS-CoV-2 samples with a cycle threshold (Ct) value of ≤32 were submitted for further analysis.

Multiplex Real-time reverse transcription polymerase chain reaction (RT-PCR): Positive specimens by routine RT-PCR, were analyzed using the RUO SARS-CoV-2 Variant Set 1 Test performed on the Cobas 6800 System (Roche, Branchburg, NJ). The variant assay is an automated, multiplex, real-time reverse transcription polymerase chain (rt-PCT) for the assessment of E484K, N501Y and del69/70 of the SARS-CoV-2 spike gene. 800 uL per specimen were submitted per the manufacturer’s instructions. Archival SARS-COV-2 samples from previously assessed samples (prior to July 2021) of varying lineages were also assessed during validation, results are below. All findings are consistent with prior NGS identification (supplemental table 1).

Supplemental table 1

| Material | NGS | SCI | E484K | N501Y | del 69-70 |
| --- | --- | --- | --- | --- | --- |
| *Archival* | Epsilon | 28.18 | - | - | - |
| *Archival* | Alpha | 22.45 | - | 25.94 | 22.09 |
| *Archival* | Beta | 21.29 | 25.82 | 24.88 | - |
| *Archival* | Delta | 28.87 | - | - | - |
| *Archival* | Iota | 27.74 | - | - | - |
| *Archival* | Mu | 27.55 | 32.37 | 31.17 | - |
| *SCI: Sample Check Indicator, NGS: Next Geneation Sequencing* | | | | | |

Viral whole genome sequencing: Nucleic acid automated extraction and Isolation occurred using the MagMAX Viral/Pathogen Ultra Nucleic Acid Isolation Kit (Thermo-Fisher, Waltham, MA) within the KingFisher Presto 96DW (Thermo-Fisher) in conjunction with the Hamilton Microlab STARPlus Instrument (Hamilton, Reno, NV). cDNA reverse transcription was carried out using Superscript III (Thermo-Fisher). All were performed using the manufacturer’s instructions. Targeted sequencing was performed using the Ion AmpliSeq SARS-CoV-2 Panel (Thermo-Fisher). Barcoded multiplexed libraries (batches of eight) were prepared with Ion 530 chips on the Ion Chef (Thermo-Fisher) and sequenced on the Ion Torrent S5 (Thermo-Fisher) according to manufacturer recommendations.

Bioinformatic Pipeline: SARS-CoV-2 reference alignment (GenBank ID: MN908947) and variant calling was performed by the Ion Torrent Suite 5.10.1 Ion Reporter. On target and uniformity coverage of 90% was required and mean depth coverage of 1000 was required for reporting. FASTA files were generated using the generateConsensus plugin available at <https://apps.thermofisher.com/apps/spa/#/publiclib/plugins>. Epidemiologic Clade and Pangolin lineage classifications were generated from these FASTA files using NextStrain.org and pangolin.cog-uk.io, respectively, the weekend of 12/18/2021.
